# Parameter Significance Test Using Mixture Models (PaSTUM) allowing Type-1 Error Control for Exposure-Response Modelling

**DOI:** 10.1101/2025.06.13.25329543

**Authors:** Daniel Wojtyniak, Jinju Guk, Sebastian G. Wicha

**Author notes:** **Corresponding author:** Prof. Dr. Sebastian G. Wicha, Professor of Clinical Pharmacy, University of Hamburg, Institute of Pharmacy, Bundesstraße 45, 20146 Hamburg, Germany.

## Abstract

In drug development, exposure-response models are widely used to inform decisions in dose optimization processes. Type I error (T1) due to mis-specified models can lead to critical and costly decisions. Therefore, a new approach called: “parameter significance test using mixture models (PaSTUM)” is proposed and compared against the standard approach (STA). A stochastic simulation estimation workstream was performed to test T1 rate, power, precision, and accuracy of drug-effect parameter estimates as well as predictive performance of the models. A total of 78 simulation scenarios for a hypothetical antidiabetic drug were investigated. For the T1 investigation, the arm allocation was randomly permutated and the AUC values were randomly sampled with replacement to each patient. For the power investigation, no arm permutation was performed, and the treatment arm remained unchanged while only the placebo patients got randomly sampled AUC values from the treatment patients. The relative root mean square error (rRMSE) and relative bias (rBias) were analyzed for predictive performance. Precision and accuracy for the drug-effect parameter were compared to the STA parameter estimates for each structural model with the most patients. PaSTUM outperformed STA regarding T1 rate inflation (20/78,78/78 > 6.53%) for PaSTUM and STA respectively. Power was marginally worse for PaSTUM. rRMSE and rBias for the full PaSTUM and STA models were similar in the power setting, but PaSTUM outperformed STA in the T1 setting. Precision and Accuracy of parameter estimates were similar for PaSTUM and STA.

## Introduction

Nonlinear mixed effects (NLME) models are a commonly used method to assess the effectiveness of drugs in clinical development [1]. These models quantify the relationship between drug treatment-, dose-, or exposure- and a certain response. Amongst those, Exposure-Response (E-R) models are the most complex ones, as they quantitively evaluate the relationship between drug exposure and a certain response. Moreover, these approaches assist in the identification of sub populations, which might require different dosing regimens to achieve similar responses [2]. The US Food and Drug Administration (FDA) [3] as well as the European Medicines Agency (EMA) [4] recommend the use of such E-R NLME models for decision making during and across clinical studies.

During model development, hypothesis tests are performed to test if an added parameter pertaining drug efficacy would improve the model performance significantly. Thereby, the likelihood ratio test is conducted, where the likelihoods of two nested models, a so-called full model and a reduced base model, are compared against each other. The test statistic is assumed to follow a chi-square distribution [5] with degrees of freedom equal to the difference of estimated parameters between the full and base model. Nevertheless, misspecifications in one, or both models as well as multiple testing can lead to parameters improving the data description, even though the parameter may capture random noise. This is referred to as a type I error (T1). The opposite can also occur, so removing/abstaining from the inclusion of a parameter even though this parameter or effect is actual present. Which is called a type II error. During clinical drug development control of both errors is crucial for adequate decision making.

To this end several different methods have been proposed and published specifically to control T1 rate [6]. Two recently introduced methods are “Individual Model Averaging” (IMA), and “randomized combined Likelihood Ratio Test-Modelling” (rcLRT-Mod) [6]. IMA utilizes mixture models to control the T1 rate. It thereby assigns patients with certain probabilities to two different sub models, a drug effect and a zero-drug effect sub model. These sub models are the same in the base and full IMA model. The difference between base and full model is how the patients from the dataset are assigned to the different sub models using the mixture. In contrast to the conventional approach, the likelihood ratio test statistics evaluates if the mixture proportion parameter between the treatment and the placebo arm can be estimated with statistical significance. Hence, in case of no apparent treatment-response, all patients can be randomly assigned to either sub model without compromising the model performance, whereas for a significant treatment-response, treatment and placebo patients have distinct probabilities to be assigned to either sub model with a significant proportion parameter. IMA thereby has shown to reduce T1 inflation due to placebo model misspecification in treatment-response settings compared to the standard approach (STA) [7]. In a different publication IMA as well as rcLRT-Mod and five other methods were compared against each other regarding T1 rate and power. There, it was shown, that IMA and rcLRT-Mod were the only two methods that controlled the T1 rate in presence of placebo model misspecification. They also indicated, that IMA outperformed rcLRT-Mod regarding power, especially with smaller drug effect sizes [6]. In a preprint of the same first author, two different methods to apply IMA with uneven placebo and treatment arm sizes (a normal IMA method and saturated IMA) were proposed. On top a method where IMA is applied in a dose-response setting was introduced. Yet this preprint did not focus on T1 rate in the dose response setting and therefore only evaluated precision and accuracy of the drug effect parameter under placebo model misspecification [8]. Moreover, IMA is not applicable in E-R without changing the methodology, as it relies on both placebo and treatment patients being assignable to the drug effect sub model. Yet, in E-R the drug effect is directly linked to the exposure and thereby placebo patients will always show a zero-drug effect even when allocated to the drug effect sub model.

The goal of the current study is to develop a method called: “Parameter Significance Test Using Mixture Models” (PaSTUM), that addresses the problems of the IMA method so that it could be applied for E-R analysis. The method is presented and compared against a normal E-R analysis regarding T1 rate, statistical power, predictive performance, as well as precision and accuracy of the estimated drug effect parameter.

## Methods

### Software

To simulate and estimate the data, NONMEM (ICON plc, Ireland, Version 7.5) as well as PsN (Uppsala University, Sweden, Version 5.3.1) was used. The visualization, calculations and automatization of the workstream was done with R (Version 4.2.0) executed via Visual Studio Code (Version 1.95).

### Study-Design

A simulation study of a hypothetical antidiabetic drug with effect on fasting plasma glucose (FPG) given via infusion was conducted to investigate the new method. The treatment arm consisted of either two or three different spaced dose groups (low, medium or high) such that the resulting area under the concentration-time curve (AUC) values do not overlap, to ensure a clear differentiation in response across different dose groups. The placebo arm was of equal size as the treatment arm. Simulations were performed with varying patient numbers of 3, 10, 30, or 100 patients per dose group, yielding a total of 16 combinations (4 different number of patients per dose group x 4 combinations of two or three low, medium, or high dose groups). To evaluate the effect of different structural models, three direct pharmacodynamic (PD) models with the following E-R relationships: linear, loglinear and emax as well as an indirect PD model, namely a turnover model with an emax effect on Kout were tested (Table 1) (Appendix 1). To ensure similar effect sizes across different E-R relationships, different doses were administered for different structural models. On top, three additional scenarios (Table 2) were simulated and evaluated.

**Table 1.**
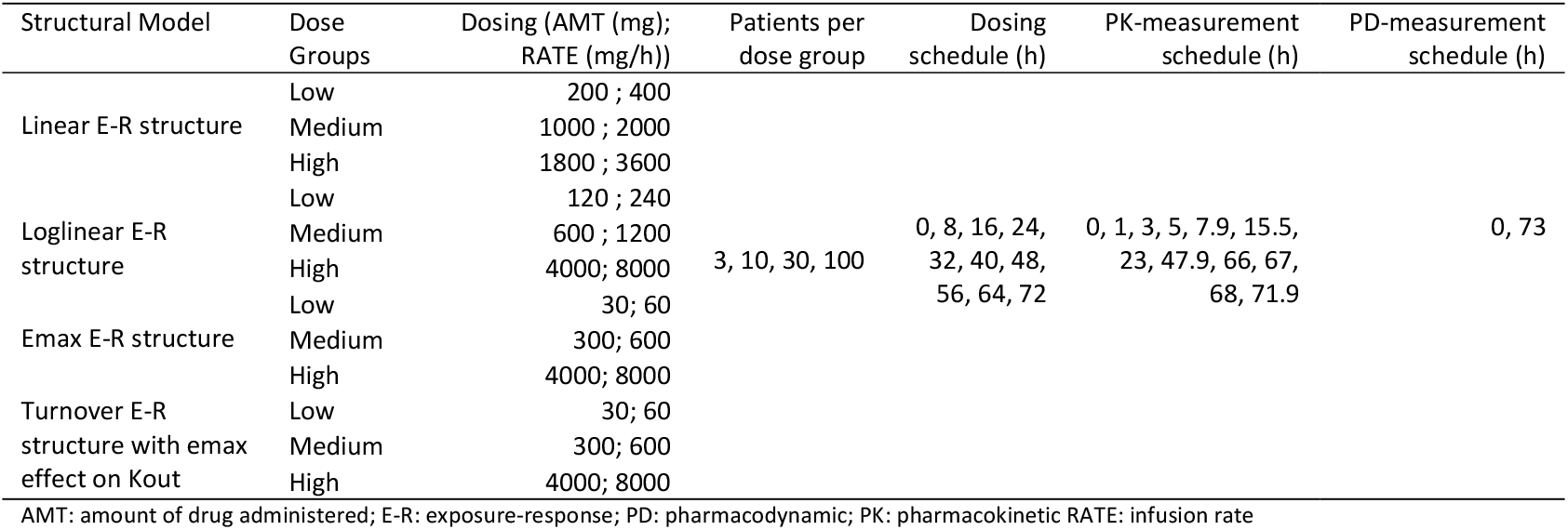
Dosing schemes as well as PK- and PD-measurement schedule for the different scenarios tested.

**Table 2.**
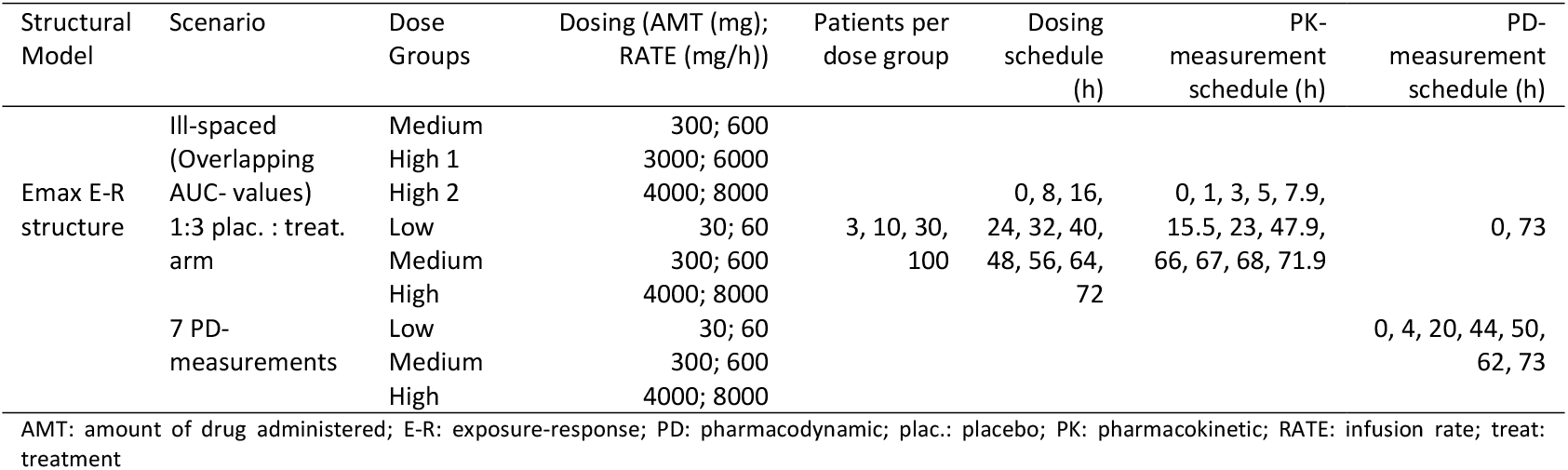
Additional scenarios tested to investigate the method behavior under different conditions.

### Standard Approach and Parameter Significance Test using Mixture Models

#### Standard Approach

For the standard approach, the base model is without drug effect parameter (Equation 1). The full model on the other hand adds one or multiple parameters describing an E-R correlation (Equation 2).

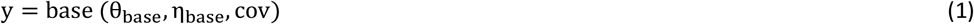

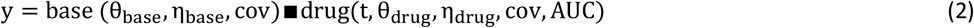

y is the individual prediction, base is a function describing the baseline observation, drug is a function describing the impact of a drug exposure, ∎represents any arithmetic operation like addition or multiplication, t is time, *θ* are fixed effects while *η* are random effects of the function, *cov* is the covariate vector and AUC is the exposure metrics for each individuum.

#### PaSTUM

PaSTUM overcomes the issue, that for E-R analysis the two sub models from the original IMA approach do not differ for the placebo arm.

To overcome this issue, in the PaSTUM approach, ‘shadow’ AUC values are randomly assigned to the placebo arm from the treatment arm by sampling with replacement. The models themselves are mixture models with two sub models: sub model 1 with a drug effect (Equation 2) and sub model 2 with a zero-drug effect (Equation 1). For the likelihood ratio test, the difference between base and full model is the proportion parameter of the mixture model assigning each subject to the different sub models. For the base model the probability is equal for each subject (Equation 3) while in the full model the probability is estimated and dependent on the arm assignment (Equation 4). For the additional scenario, where the treatment and placebo arm are of different size, the equation for the base (Equation 5) and full (Equation 6) PaSTUM models are changed to mimic the saturated IMA structure [8].

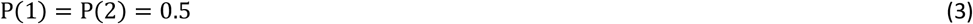

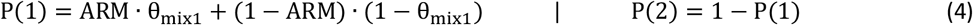

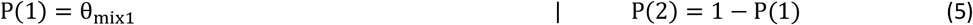

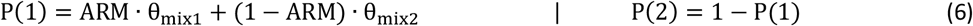

ARM is either 0 for placebo patients or 1 for treatment patients, *θ*_mix_ are the estimated mixture proportion parameter ∈ [0,1].

Both methods compare the benefit of adding one or multiple parameters between base and full model using a likelihood ratio test. For STA the degrees of freedom equal the number of drug effect parameter added, while for PaSTUM the degree of freedom is one, for the only added parameter is the added mixture proportion parameter. Code examples for both methods are given in the supplemental material (Appendix 1).

### Simulation and Estimation Workstream

The base analysis workstream for the stochastic simulation and estimation (SSE) analysis was the following: Simulation of a new dataset using a Pharmacokinetic (PK)-PD model utilizing the original dataset, followed by an estimation step in which the AUCs are estimated using the same structural PK model, which are in turn used for the E-R analysis.

For the T1 investigation (no real drug effect simulated) the arm allocation is randomly permutated. Then, the AUC values from the treatment arm are sampled with replacement for all patients, respectively. For the power investigation (real drug effect simulated) no permutation is performed and only the placebo arm is assigned sampled AUC values from the treatment arm, while the treatment arm itself remained unchanged. After that, the different models for STA and PaSTUM were used to analyze the data. Afterwards, the respective full and base models were compared regarding their objective function value (OFV).

### Analysis of Type I error rate

To investigate the T1 rate, 1000 simulation were conducted following the previously mentioned workstream for the T1 rate. The total number of times the respective full model was considered better than the respective base model divided by the total number of simulations and estimations that resulted in a succeeded NONMEM execution (Appendix 2) is the T1 rate (Equation 7). Following a binomial distribution, the 95% confidence interval (CI) for a 5% T1 rate after 1000 simulations is between 3.81% and 6.53% (Equation 8). Therefore, every T1 rate above 6.54% will be seen as inflated.

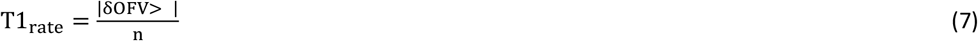

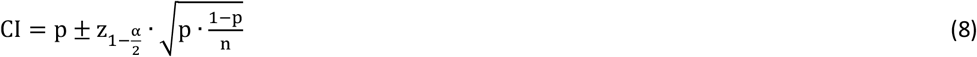

Where || counts the scenarios for which the criteria inside are met, δOFV is the difference in OFV between full model and base model, C is the critical value following a chi-square distribution with degrees of freedom equal to the number of drug effect parameter for STA and one for PaSTUM, n is the total number of succeeded runs, CI is the 95% CI, p is the expected proportion of interest (in this analysis always 0.05), 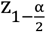 is the z-value for the desired level of confidence (in this analysis always 95%).

### Analysis of Power

For the analysis of power, 1000 simulations, following the workstream with a simulated drug effect present, were conducted. The total number of times the respective full model outperformed the respective base model divided by the total number of simulations and successful estimations is the power or one minus the type II error (Equation 9). The power was considered acceptable if it was above 80% in this analysis.

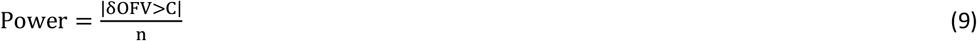

### Analysis of Predictive Performance

For the investigation of the predictive performance the mean of the relative root mean square error (rRMSE) and its 95% CI as well as the mean of the relative bias (rBias) and its 95% CI of 1000 SSEs was calculated for each base and full model respectively, comparing the difference between simulated and model-predicted FPG values (Equation 10-11). rRMSE values are compared within each setting, where the lower the rRMSE value the better the model description for the individual scenario. For the rBias, the success criterion was defined to be the inclusion of zero in the 95% CI, as this would mean zero bias was included in the 95% CI for the corresponding model prediction.

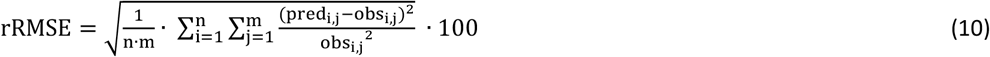

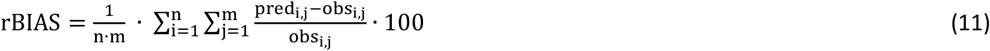

where n is the total number of individuals, m is the total number of observations per individual, predi,j is the prediction for FPG for the ith individual at the jth time point, obs_i,j_ is the observed FPG for the ith individual at the jth time point.

### Analysis of Precision and Accuracy of E-R parameters

To investigate the precision and accuracy for the drug effect parameters, 1000 simulations under the power setting were investigated regarding their estimations for the drug effect parameters for each base and full model respectively. The mean and 95% CI of the estimated parameters for each different scenario were then compared to the STA full model scenario with 100 patients per dose group and three dose groups for the respective drug effect structure, as this estimated drug effect parameter was considered the “true” parameter (Equation 12–14). The success criterion was met, if the 95% CI overlapped with the “true” parameter.

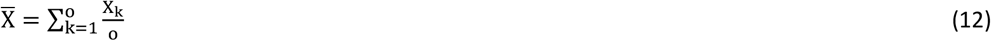

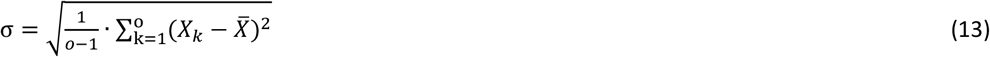

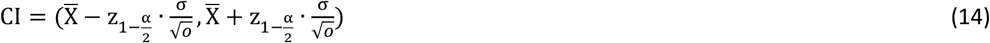

Where 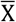 is the mean of a predicted effect parameter for o simulations for a specific setting, X_k_ is the estimation for the kth simulation, o is the total number of simulations for each setting, σ is the standard deviation of a predicted effect parameter for o simulations for a specific setting.

## Results

### Type I Error and Power

Table 3 presents the T1 rate and Table 4 the statistical power for PaSTUM and STA under different scenarios. The T1 rate for PaSTUM was controlled in most cases (PaSTUM: T1 < 6.54%, 48/64), whereas it was always inflated for STA (STA: T1 < 6.54%, 0/64). Simultaneously, the uncalibrated statistical power exceeded 80% in all instances for PaSTUM (PaSTUM: Power > 80%, 64/64) and for STA (STA: Power > 80%, 64/64).

**Table 3:** Type I error rate for different SSE Scenarios (runs which achieved minimization successful)

| Setting | PaSTUM |  |  |  | STA |  |  |  |
| --- | --- | --- | --- | --- | --- | --- | --- | --- |
|  | linear | loglinear | emax | turnover | linear | loglinear | emax | turnover |
| L-H-3 | 9.97% | 3.94% | 8.64% | 9.65% | 46.2% | 12.5% | 45.3% | 23.0% |
| L-H-10 | 6.40% | 4.46% | 4.99% | 5.23% | 64.5% | 27.0% | 73.2% | 53.5% |
| L-H-30 | 6.74% | 2.83% | 4.21% | 5.50% | 81.6% | 80.2% | 95.8% | 91.7% |
| L-H-100 | 7.01% | 2.40% | 2.50% | 10.6% | 95.3% | 100% | 99.7% | 99.1% |
| L-M-3 | 9.27% | 5.49% | 7.30% | 8.43% | 38.6% | 16.0% | 30.8% | 22.8% |
| L-M-10 | 6.18% | 5.48% | 6.35% | 6.64% | 60.9% | 41.8% | 57.8% | 54.7% |
| L-M-30 | 5.90% | 3.07% | 4.70% | 5.58% | 87.2% | 92.2% | 90.3% | 97.1% |
| L-M-100 | 5.40% | 4.00% | 5.20% | 6.28% | 99.5% | 100% | 100% | 99.9% |
| M-H-3 | 7.40% | 4.42% | 7.20% | 8.42% | 37.9% | 16.5% | 37.7% | 18.4% |
| M-H-10 | 4.51% | 3.75% | 5.71% | 4.47% | 48.5% | 48.7% | 59.5% | 54.5% |
| M-H-30 | 4.91% | 4.20% | 5.21% | 5.69% | 42.6% | 95.9% | 80.1% | 96.3% |
| M-H-100 | 5.11% | 4.40% | 5.50% | 6.11% | 18.4% | 100% | 91.0% | 100% |
| L-M-H-3 | 5.70% | 4.31% | 6.81% | 6.64% | 47.8% | 15.4% | 52.6% | 28.3% |
| L-M-H-10 | 5.48% | 2.53% | 5.65% | 6.11% | 67.0% | 49.3% | 78.1% | 71.6% |
| L-M-H-30 | 5.02% | 1.80% | 3.40% | 5.03% | 76.7% | 97.4% | 95.9% | 98.8% |
| L-M-H-100 | 5.81% | 3.40% | 5.40% | 7.07% | 76.2% | 100% | 99.7% | 98.4% |

H: high dose, L: low dose, M: medium dose, PaSTUM: Parameter significance test using mixture models, STA: standard approach, blue: T1 0-3.72%, green: T1 3.81-6.53%, red: T1 50-100%, yellow: T1 6.55-49.9%

**Table 4:** Power for different SSE Scenarios (runs which achieved minimization successful)

| Setting | PaSTUM |  |  |  | STA |  |  |  |
| --- | --- | --- | --- | --- | --- | --- | --- | --- |
|  | linear | loglinear | emax | turnover | linear | loglinear | emax | turnover |
| L-H-3 | 98.5% | 94.7% | 95.6% | 97.8% | 100% | 100% | 100% | 100% |
| L-H-10 | 100% | 100% | 100% | 100% | 100% | 100% | 100% | 100% |
| L-H-30 | 100% | 100% | 100% | 100% | 100% | 100% | 100% | 100% |
| L-H-100 | 100% | 100% | 100% | 100% | 100% | 100% | 100% | 100% |
| L-M-3 | 97.9% | 96.7% | 89.8% | 97.9% | 100% | 100% | 100% | 99.8% |
| L-M-10 | 100% | 100% | 100% | 100% | 100% | 100% | 100% | 100% |
| L-M-30 | 100% | 100% | 100% | 100% | 100% | 100% | 100% | 100% |
| L-M-100 | 100% | 100% | 100% | 100% | 100% | 100% | 100% | 100% |
| M-H-3 | 100% | 98.5% | 100% | 99.9% | 100% | 100% | 100% | 100% |
| M-H-10 | 100% | 100% | 100% | 100% | 100% | 100% | 100% | 100% |
| M-H-30 | 100% | 100% | 100% | 100% | 100% | 100% | 100% | 100% |
| M-H-100 | 100% | 100% | 100% | 100% | 100% | 100% | 100% | 100% |
| L-M-H-3 | 100% | 99.3% | 100% | 100% | 100% | 100% | 100% | 100% |
| L-M-H-10 | 100% | 100% | 100% | 100% | 100% | 100% | 100% | 100% |
| L-M-H-30 | 100% | 100% | 100% | 100% | 100% | 100% | 100% | 100% |
| L-M-H-100 | 100% | 100% | 100% | 100% | 100% | 100% | 100% | 100% |

H: high dose, L: low dose, M: medium dose, PaSTUM: Parameter significance test using mixture models, STA: standard approach, green: Power 80.0-100%, red: T1 0-79.9%

### Predictive Performance

Figure 1 and Figure 2 display the mean rRMSE and rBias for the observed vs. model-predicted FPG, along with their 95% CIs across all scenarios and models. The base STA model showed the worst performance in all scenarios, regardless of the setting. While the full STA model performed poorly only under the T1 setting, it still displays better results compared to the base STA model. In contrast, both the base and full PaSTUM models consistently outperformed the STA models or performed equally well. Generally, the full models outperformed the corresponding base models across most scenarios. The 95% CI for the rBias of PaSTUM typically included or was close to 0, indicating that these models were unbiased or only slightly biased with 95% confidence. Contrariwise, the 95% CI for the base STA model frequently excluded 0, suggesting bias in this model, while the full STA model showed similar unbiased results as the PaSTUM models. Overall, the log-linear models performed worse compared to all other structural models.

**Figure 1.**
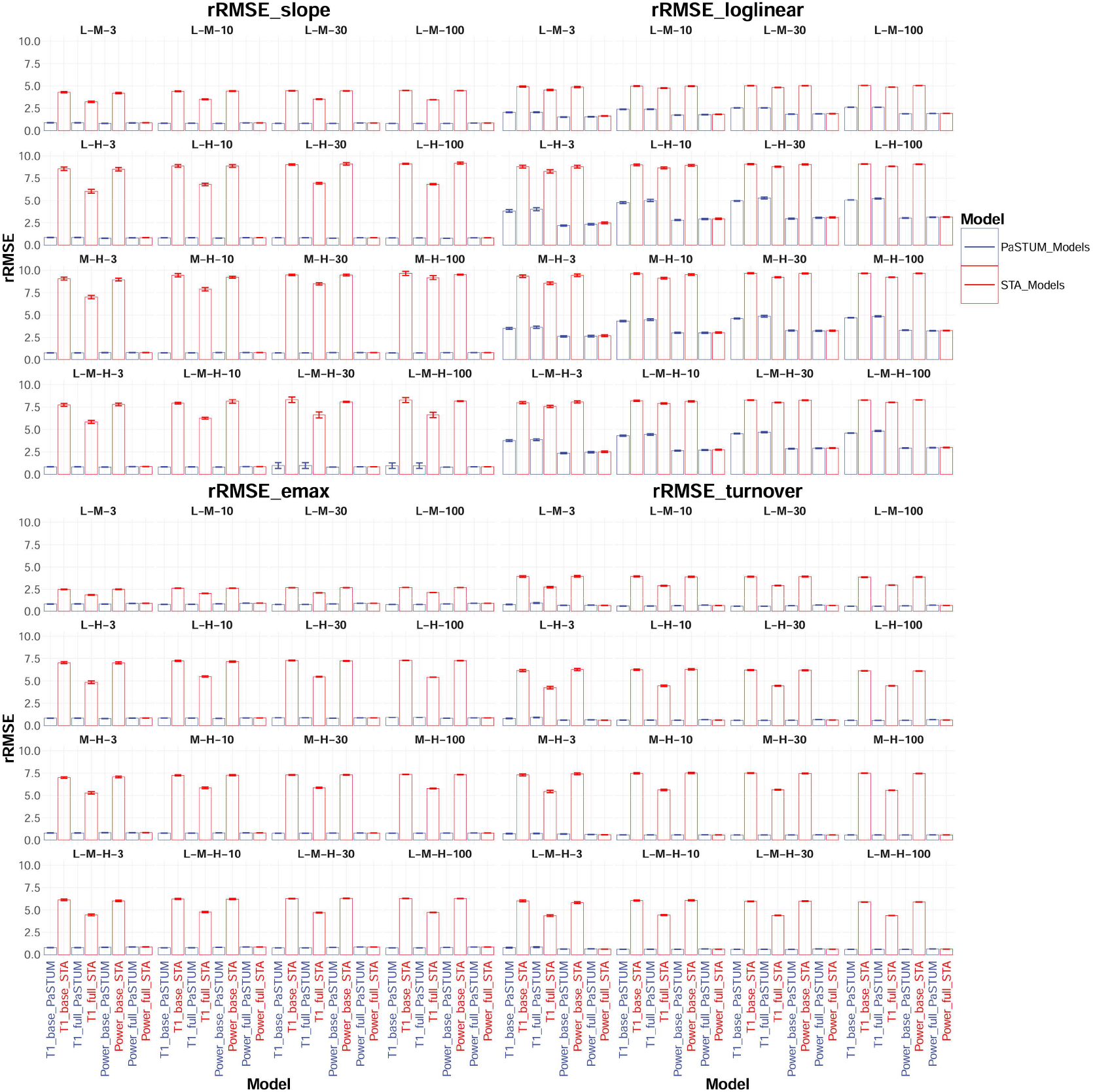
Mean rRMSE and 95% confidence intervals for all models and for all different SSE scenarios

**Figure 2.**
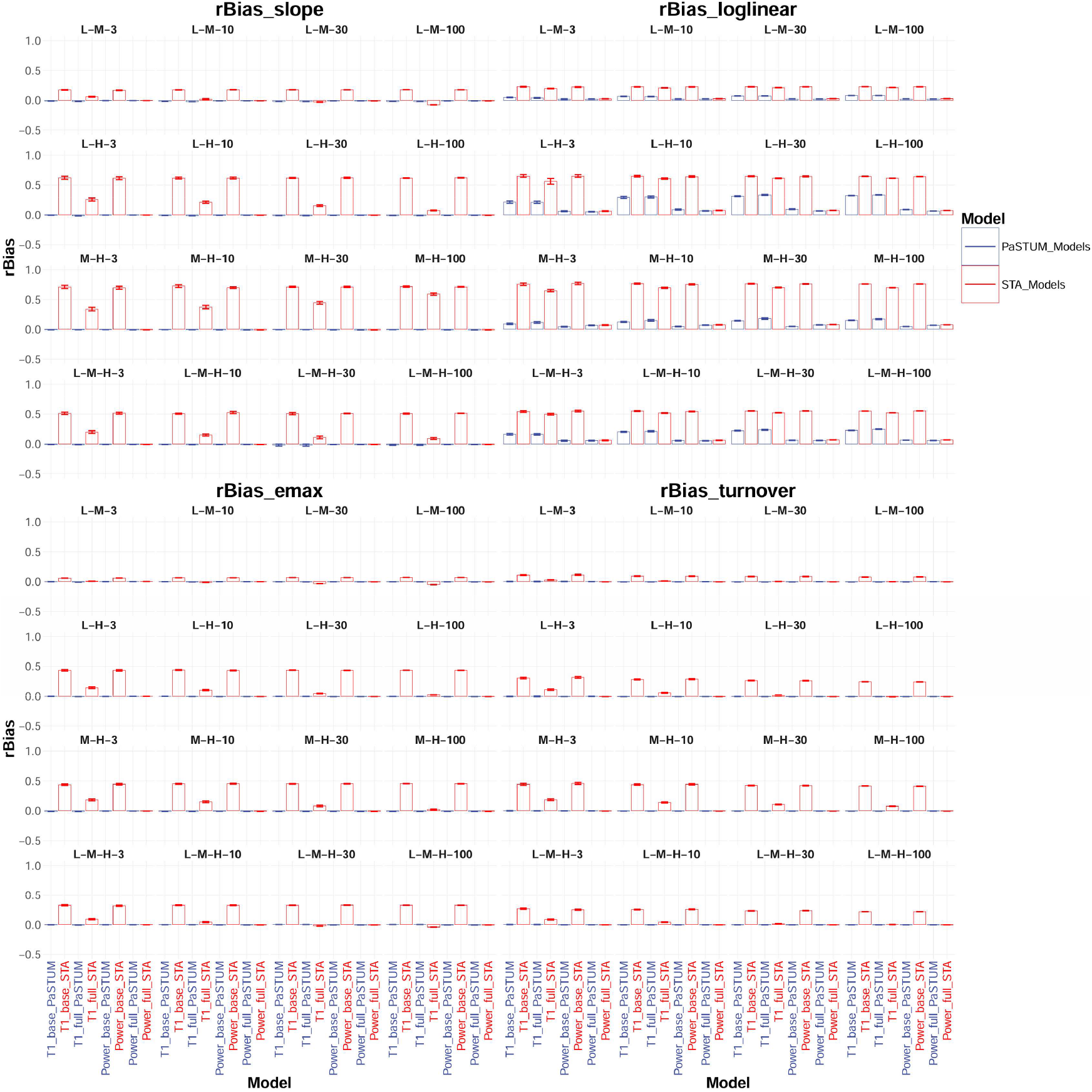
Mean rBias and 95% confidence intervals for all models and for all different SSE scenarios

### Precision and Accuracy of E-R Parameter

The precision and accuracy of the drug effect parameter estimates improved with an increasing number of patients per group. The baseline FPG parameter was estimated with sufficient precision across most scenarios and models, except for the base STA models, where the baseline parameter was regularly underestimated. The precision and accuracy of the remaining effect parameters varied by scenario and model. For more complex drug effect structures, such as indirect turnover with emax effect on Kout and direct emax models, the drug effect parameter (baseline FPG, EC50 and EMAX), were often estimated with lower precision and higher variability within a given setting compared to the parameter for simpler drug effect structures like direct linear or log-linear models (baseline FPG and slope). Figure 3 presents the mean predictions along with their 95% CIs. For interpretability, estimates differing from the “true” value by more than a five-fold difference for the Emax and turnover models or a 0.2-fold difference for the linear and log-linear models were excluded from the plots.

**Figure 3.**
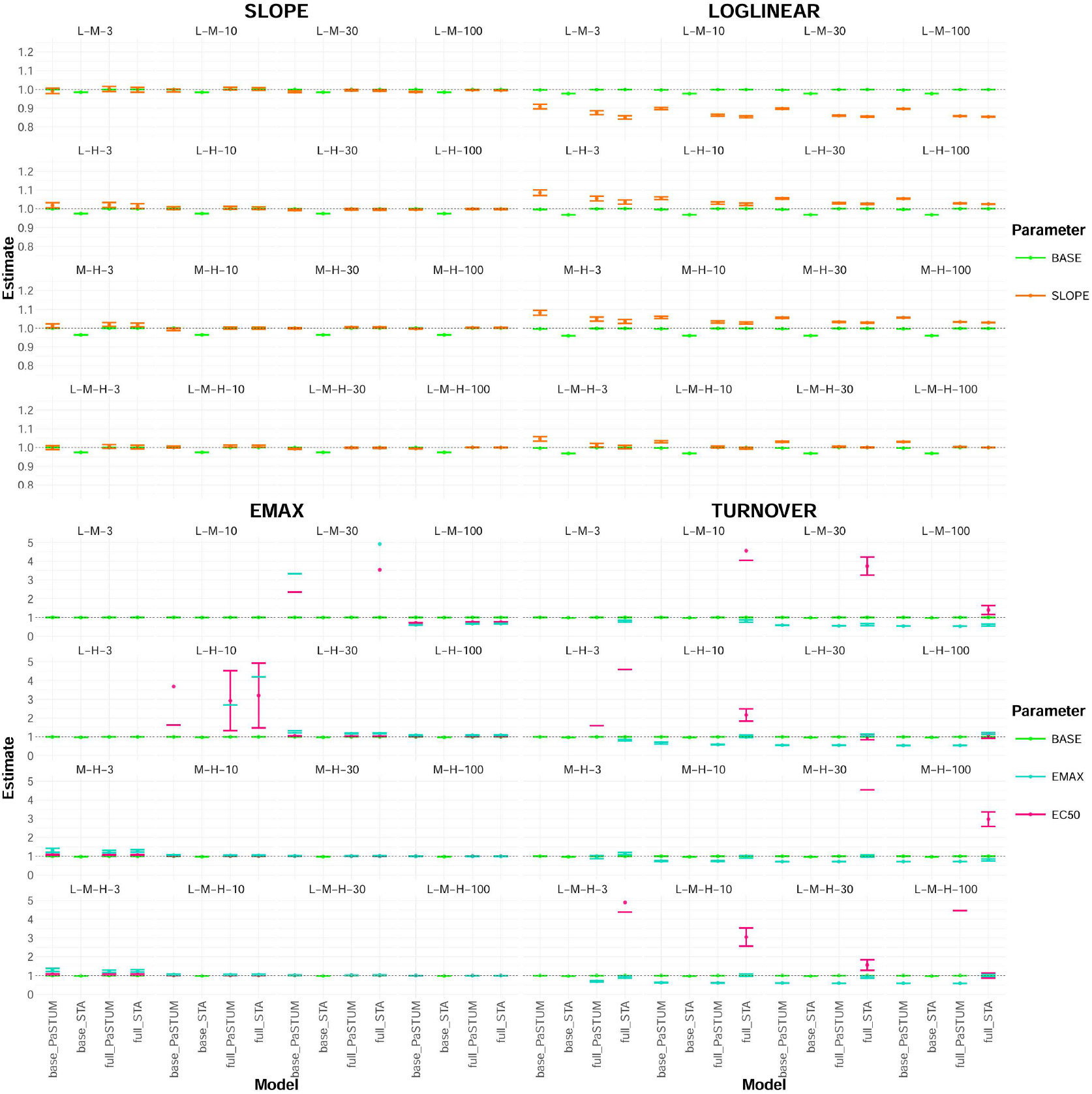
Relative Parameter mean estimates and 95% confidence intervals under the power Setting (estimates outside a 0.2-fold for slope and loglinear and a 5-fold range for emax and turnover model are excluded for readability purposes)

### Additional Scenarios

#### Type I Error and Power

The results for the T1 rate and Power in the three additional scenarios are summarized in Table 5. Overall, the T1 rate for PaSTUM was better controlled than for STA. Except for the scenarios with multiple PD measurements and those with only nine patients in total, the T1 rate for PaSTUM remained controlled (PaSTUM T1 > 6.54%: 4/12). In contrast, the T1 rate was inflated across all scenarios for STA (STA T1 > 6.54%: 12/12). The Power exceeded the 80% threshold in all simulated scenarios for both STA and PaSTUM.

**Table 5:**
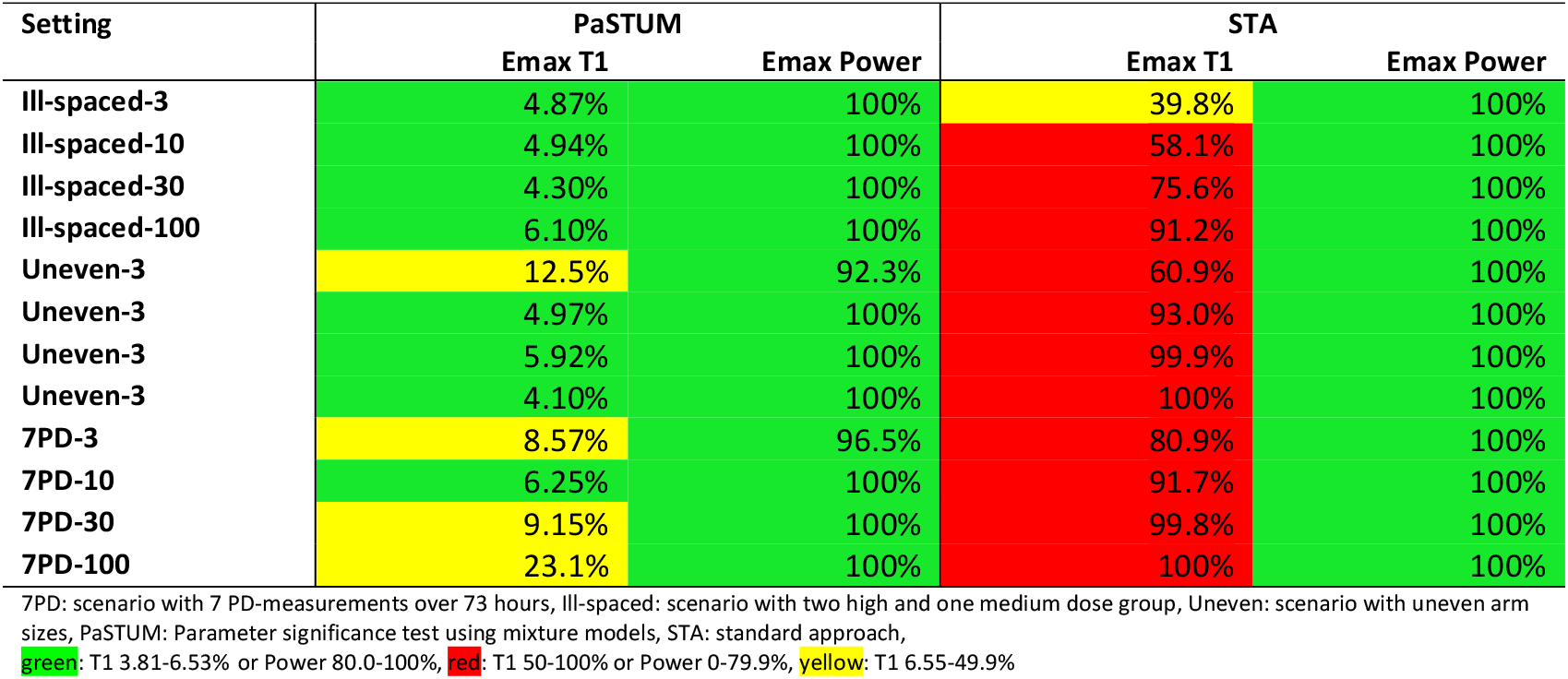
Type I error rate and Power for additional SSE Scenarios (runs which achieved minimization successful)

### Predictive Performance

The mean as well as 95% CI for rRMSE and rBias for the additional scenarios follow a similar trend to the main analysis as shown in Figure 4. The base STA model was the worst performing model, while the full STA model also described the data poorly in the T1 setting. PaSTUM base and full model on the other hand described the data good in all settings and were less often biased.

**Figure 4.**
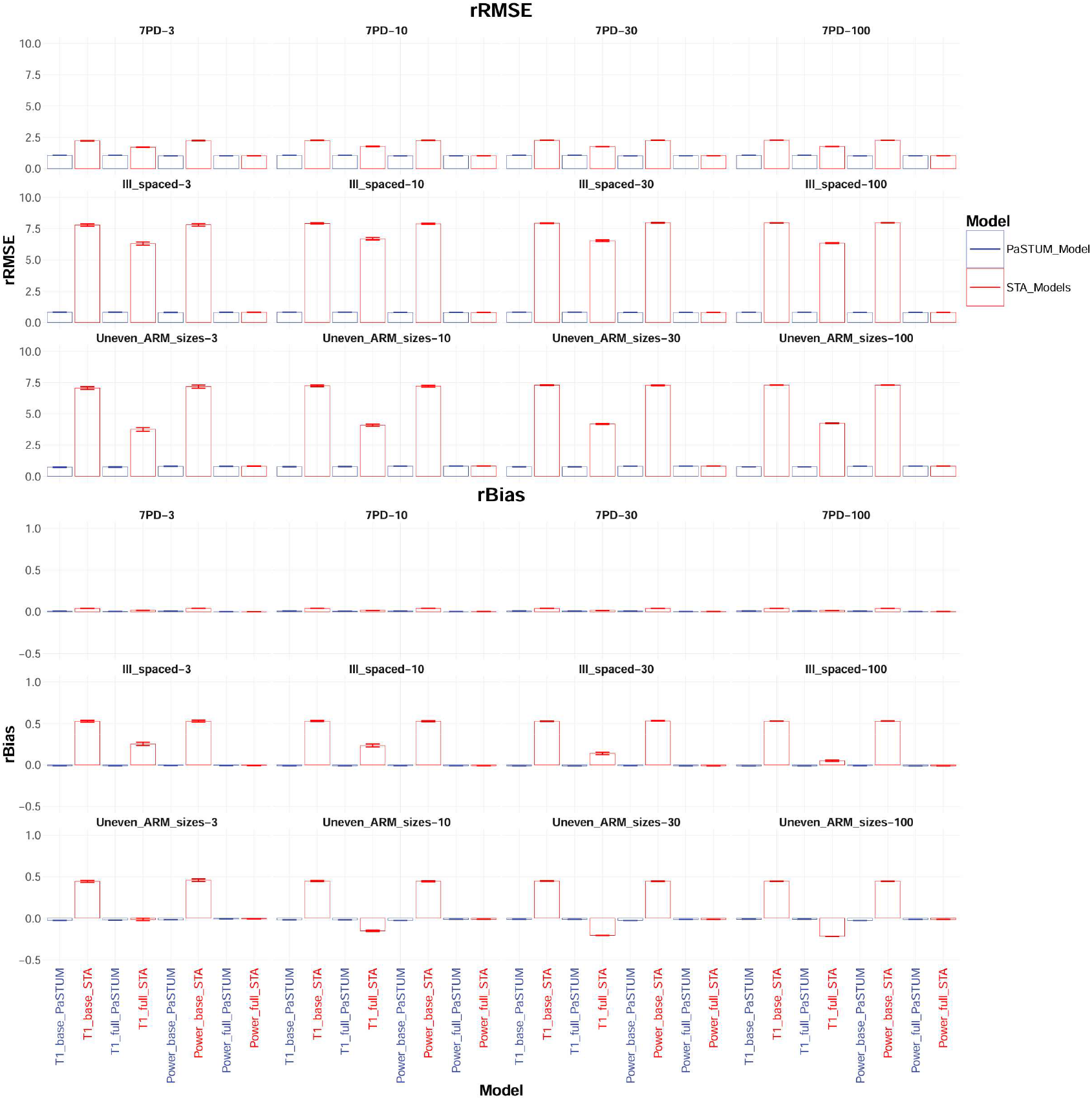
Mean and 95% confidence intervals for the rRMSE and rBias for all models and for all different additional SSE scenarios

### Precision and Accuracy of E-R Parameter

The prediction of the drug effect parameters was in the additional scenarios similarly good as in the other scenarios as shown in Figure 5. The drug effect parameters were estimated with lower bias and higher precision with more patients. In the scenario with multiple PD measurements the drug effect parameters were underestimated by roughly 75% for Emax and 50% for EC50. Overall, there were no significant differences between the estimation of the drug effect parameter for all different methods in the models, with exception of the base-STA model, which underestimated the baseline FPG level in all scenarios.

**Figure 5.**
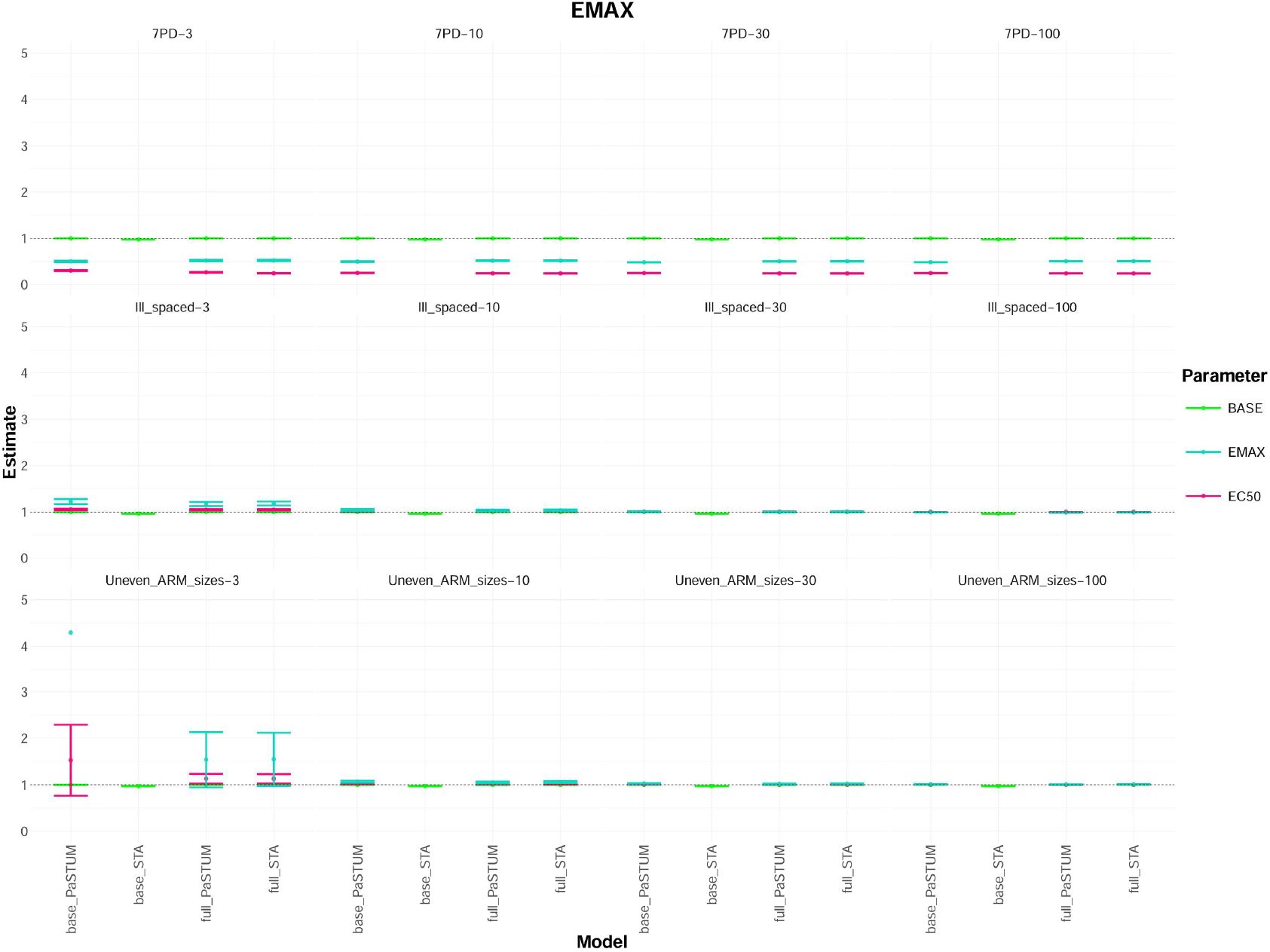
Relative Parameter mean estimates and 95% confidence intervals for all different additional SSE scenarios under the power Setting (estimates outside a 5-fold range are excluded for readability purposes)

## Discussion

This paper introduced the novel PaSTUM method to control T1 rates for E-R modelling. The method represents an evolution of the IMA approach [7] utilizing mixture models for significance testing and sampling approaches to handle exposure variables in the control arm. The PaSTUM method was comprehensively evaluated and compared to STA.

Overall, PaSTUM consistently outperformed STA in all tested scenarios with respect to T1 rate. Instances where the T1 rate was inflated for PaSTUM occurred in scenarios, where inflated T1 rate is expected, like scenarios with a low number of patients or where the predictive performance of all models was inherently low. Even in these scenarios, PaSTUM T1 rate is not nearly as inflated as STA T1 rate. While STA demonstrated slightly better power in some scenarios, this difference was marginal. Across the 78 scenarios tested (64 main analysis and 12 additional scenarios), the power for PaSTUM was below 80% not even once. Thus, regarding statistical power, PaSTUM performed comparably to STA with only slight differences in extreme cases.

As the T1 rate for PaSTUM got inflated especially in the scenarios with small sample sizes, i.e. a low number of patients, a different variation of the method was considered to maybe hold better results. The hypothesis hereby was, that the number of AUCs to resample may have been too small. Therefore, a variation, where instead of randomly sampling AUCs from the treatment arm, the placebo patients got imputed the geometric mean of all treatment patient AUCs instead. The geometric mean was chosen because AUCs are assumed to be log-normally distributed across patients [9]. Indeed, this approach showed less inflated T1 rates for small sample sizes in some scenarios, however the T1 rate was more and severely inflated in some other scenarios. The complete comparison between both variations of PaSTUM is shown in Appendix 3

The predictive performance of FPG levels was equivalent or better for PaSTUM as for STA. In fact, in scenarios where no true drug effect was present, PaSTUM provided FPG estimates closer to the simulated values than those from STA. Parameter estimation was also robust for PaSTUM, with base and full models frequently including the “true” parameter values within their 95% CIs, while especially the base STA model consistently underestimated the baseline FPG value. It is worth noting that the “true” parameter values were defined as the estimates derived from the full STA model in a setting with 100 patients per dose group and three dose groups (600 patients in total), thus, no model could surpass STA in parameter estimation under these conditions.

In additional scenarios involving multiple PD measurements within 73 hours, an unevenly spaced dosing design (with two doses near the maximum effect size and overlapping AUC values), and unequal sample sizes between the placebo and treatment arms, results were largely consistent with the other scenarios. However, in the case of multiple PD measurements, none of the models precisely estimated the drug effect parameter, likely due to an uninformed study design.

In terms of runtime and stability, PaSTUM is inferior to STA. Due to the mixture model property, PaSTUM takes a longer run time and tends to be less stable than STA. While this is manageable for simpler models with limited data, more complex models with larger datasets may require several days to complete using PaSTUM. Stability-wise, PaSTUM is also more susceptible to early termination before finalizing the minimization process (Appendix 2). However, in none of the tested scenarios did STA or PaSTUM failed in the minimization process entirely.

The estimation process for PaSTUM is more sensitive to poorly chosen initial estimates than STA. When initial estimates deviated significantly from the true values, the mixture model may incorrectly assign patients to sub-models, resulting in inaccurate estimates of drug effect size and, in the worst case, inflation of the T1 rate. To mitigate this risk, it is recommended to test several different initial estimates when applying PaSTUM, which in term prolongs the time needed even further.

In addition to PaSTUM, several other methods have been proposed to control the T1 rate, including IMA and rcLRT-Mod. IMA has been demonstrated to control the T1 rate in treatment-response settings across various studies [6–8]. However, as it misses the randomly sampling of dosing or exposure information to placebo patients, it is inapplicable in E-R or dose-response settings. Although the original first author published a preprint applying a modified version of IMA to a dose-response scenario, they did not investigate the T1 rate in this context [8].

The development of PaSTUM was motivated by the wish to adapt the principles of the IMA method for E-R settings while maintaining the T1 rate control observed in treatment-response scenarios. However, the required methodological adjustments led to the creation of a novel approach. Similarly, rcLRT-Mod has demonstrated reduced T1 rates comparable to IMA but has been evaluated only in treatment-response scenarios [6]. Unlike IMA, rcLRT-Mod does not rely on mixture models, making it potentially also applicable to E-R settings, as it does not require all patients to influence a mixture proportion parameter. Nevertheless, in a study comparing model averaging techniques, rcLRT-Mod achieved controlled T1 rates but showed insufficient power when the drug effect was small [6].

There are some limitations to the current study. PaSTUM has only been evaluated in an SSE setting where the “true” structural model matches the estimation model. If the structural model used for estimation differs from the true structural model, the T1 rate could become inflated due to the inherent instability of the method and its reliance on well-chosen initial estimates. Moreover, real clinical data often introduces additional complexities, such as inconsistencies, missing observations, covariates, or other factors that exacerbate model instability. These challenges increase the likelihood of PaSTUM failing to estimate parameters precisely.

Additionally, this simulation study focused exclusively on scenarios without an estimated placebo effect. In the simple models and datasets tested, a placebo effect would compensate for a zero-drug effect to such a degree that neither PaSTUM nor STA would show any T1 rate nor sufficient power in any of the tested scenarios. Investigating such cases would require datasets with more than two PD measurements.

Therefore, next steps for evaluation and further improvement of the PaSTUM method include applying the method on real clinical datasets, investigating how wrongly chosen structural models influence the T1 rate and analyzing data with enough data points to also include a placebo effect. An approach to further enhance PaSTUM could be to link it to other ways to control T1 rate. For example, model averaging methods.

## Conclusion

All in all, PaSTUM offers an excellent way to control T1 rate in E-R settings while maintaining sufficient power and predictive performance as well as precise and accurate estimates for the investigated parameter.

## Data Availability

All data produced in the present study are available upon reasonable request to the authors

## Acknowledgments

Special thanks to Christoph Pfaffendorf for the discussion of scientific ideas.

## Author contributions

D.W. J.G. and S.W. conceived the presented idea and method.

D.W. performed the research.

All authors discussed the results, contributed and accepted to the final manuscript.

